# FebriDx point-of-care test in patients with suspected COVID-19: a systematic review and individual patient data meta-analysis of diagnostic test accuracy studies

**DOI:** 10.1101/2020.10.15.20213108

**Authors:** Samuel G. Urwin, B. Clare Lendrem, Jana Suklan, Kile Green, Sara Graziadio, Peter Buckle, Paul M. Dark, Adam L. Gordon, Daniel S. Lasserson, Brian Nicholson, D. Ashley Price, Charles Reynard, Mark H. Wilcox, Gail Hayward, Graham Prestwich, Valerie Tate, Tristan W. Clark, Raja V. Reddy, Hamish Houston, Ankur Gupta-Wright, Laurence John, Richard Body, A. Joy Allen, on behalf of the CONDOR steering group

## Abstract

**Background:** We conducted a systematic review and individual patient data (IPD) meta-analysis to evaluate the diagnostic accuracy of a commercial point-of-care test, the FebriDx lateral flow device (LFD), in adult patients with suspected COVID-19. The FebriDx LFD is designed to distinguish between viral and bacterial respiratory infection.

**Methods:** We searched MEDLINE, EMBASE, PubMed, Google Scholar, LitCovid, ClinicalTrials.gov and preprint servers on the 13^th^ of January 2021 to identify studies reporting diagnostic accuracy of FebriDx (myxovirus resistance protein A component) versus real time reverse transcriptase polymerase chain reaction (RT-PCR) testing for SARS-CoV-2 in adult patients suspected of COVID-19. IPD were sought from studies meeting the eligibility criteria. Studies were screened for risk of bias using the QUADAS-2 tool. A bivariate linear mixed model was fitted to the data to obtain a pooled estimate of sensitivity and specificity with 95% confidence intervals (95% CIs). A summary receiver operating characteristic (SROC) curve of the model was constructed. A sub-group analysis was performed by meta-regression using the same modelling approach to compare pooled estimates of sensitivity and specificity between patients with a symptom duration of 0 to 7 days and >7 days, and patients aged between 16 to 73 years and >73 years.

**Results:** Ten studies were screened, and three studies with a total of 1481 patients receiving hospital care were included. FebriDx produced an estimated pooled sensitivity of 0.911 (95% CI: 0.855-0.946) and specificity of 0.868 (95% CI: 0.802-0.915) compared to RT-PCR. There were no significant differences between the sub-groups of 0 to 7 days and >7 days in estimated pooled sensitivity (p = 0.473) or specificity (p = 0.853). There were also no significant differences between the sub-groups of 16 to 73 years of age and >73 years of age in estimated pooled sensitivity (p = 0.946) or specificity (p = 0.486).

**Conclusions:** Based on the results of three studies, the FebriDx LFD had high diagnostic accuracy for COVID-19 in a hospital setting, however, the pooled estimates of sensitivity and specificity should be interpreted with caution due to the small number of studies included, risk of bias, and inconsistent reference standards. Further research is required to confirm these findings, and determine how FebriDx would perform in different healthcare settings and patient populations.

**Trial registration:** This study was conducted at pace as part of the COVID-19 National Diagnostic Research and Evaluation Platform (CONDOR) national test evaluation programme (https://www.condor-platform.org), and as a result, no protocol was developed, and the study was not registered.

**Lay summary:** Tests to diagnose COVID-19 are crucial to help control the spread of the disease and to guide treatment. Over the last few months, tests have been developed to diagnose COVID-19 either by detecting the presence of the virus or by detecting specific markers linked to the virus being active in the body. These tests use complex machines in laboratories accepting samples from large geographical areas. Sometimes it takes days for test results to come back. So, to reduce the wait for results, new portable tests are being developed. These ‘point-of-care (POC)’ tests are designed to work close to where patients require assessment and care such as hospital emergency departments, GP surgeries or care homes. For these new POC tests to be useful, they should ideally be as good as standard laboratory tests.

In this study we looked at published research into a new test called FebriDx. FebriDx is a POC test that detects the body’s response to infection, and is claimed to be able to detect the presence of any viral infection, including infections due to the SARS-CoV-2 virus which causes COVID-19, as well as bacterial infections which can have similar symptoms. The FebriDx result was compared with standard laboratory tests for COVID-19 performed on the same patient’s throat and nose swab sample. We were able to analyse data from three studies with a total of 1481 adult patients who were receiving hospital care with symptoms of COVID-19 during the UK pandemic. Approximately one fifth of the patients were diagnosed as positive for SARS-CoV-2 virus using standard laboratory tests for COVID-19.

Our analysis demonstrated that FebriDx correctly identified 91 out of 100 patients who had COVID-19 according to the standard laboratory test. FebriDx also correctly identified 87 out of 100 patients who did not have COVID-19 according to the standard laboratory test. These results have important implications for how these tests could be used. As there were slightly fewer FebriDx false results when the results of the standard laboratory test were positive (9 out of 100) than when the results of the standard laboratory test were negative (13 out of 100), we can have slightly more confidence in a positive test result using FebriDx than a negative FebriDx result.

Overall, we have shown that the FebriDx POC test performed well during the UK COVID-19 pandemic when compared with laboratory tests, especially when COVID-19 was indicated. For the future, this means that the FebriDx POC test might be helpful in making a quick clinical decision on whether to isolate a patient with COVID-19-like symptoms arriving in a busy emergency department. However, our results indicate it would not completely replace the need to conduct a laboratory test in certain cases to confirm COVID-19.

There are limitations to our findings. For example, we do not know if FebriDx will work in a similar way with patients in different settings such as in the community or care homes. Similarly, we do not know whether other viral and bacterial infections which cause similar COVID-19 symptoms, and are more common in the autumn and winter months, could influence the FebriDx test accuracy. Our findings are also only based on three studies.

## Background

The global severe acute respiratory syndrome coronavirus 2 (SARS-CoV-2) pandemic (1) has put considerable pressure on health and care services worldwide. Health and care providers require diagnostic strategies to rapidly identify patients infected with SARS-CoV-2 to implement accurate segregation of patients in health and care facilities, and to ensure early administration of evidence-based therapies to patients with coronavirus disease 2019 (COVID-19). The risks of nosocomial infection are high (2) and mechanisms to ensure that the transmission of SARS-CoV-2 is limited within hospitals, care facilities, and the community are an urgent priority.

There has been rapid development of novel clinical tests to support screening and diagnosis in both symptomatic and asymptomatic patients. In particular, there have been a number of molecular, antigen, and antibody tests which manufacturers have developed for use at the point-of-care (POC) (3). In the Emergency Department setting, for example, a rapid COVID-19 test result could aid triage of the patient into the appropriate COVID-19/non-COVID-19 sections of the hospital. This may contribute to a reduction in nosocomial infection, providing significant benefits to patient pathways, workflows, and outcomes (4, 5).

There is limited published evidence on the diagnostic characteristics, reliability, and usability of many of the available POC tests for SARS-CoV-2, in particular, when used across clinical settings with varying disease prevalence. Some pre-existing POC tests may have a role in the management of patients with suspected COVID-19. The FebriDx lateral flow device (LFD) (Lumos Diagnostics, Sarasota, Florida, USA) is a CE-marked POC test that detects two host response proteins, myxovirus resistance protein A (MxA) and C reactive protein (CRP), in fingerstick blood samples. This combination of MxA and CRP is designed to distinguish between viral and bacterial respiratory infection, respectively (6-8), and therefore the test is not specific for SARS-CoV-2. MxA is an intracellular protein that is exclusively induced by type I interferon (IFN) as part of the antiviral host response but not by other cytokines expressed during bacterial infection (9, 10). Type I IFNs are produced in response to a wide range of viral infections and are found to be elevated in the presence of most acute viral infections (7), therefore providing strong theoretical grounds to expect a rise in MxA in response to SARS-CoV-2 infection. However, a raised MxA level is not diagnostic of SARS-CoV-2 due to it being a non-specific immune response to a number of respiratory infections. Thus, the optimal use case of FebriDx is unlikely to be for ‘ruling in’ COVID-19, although it may have utility in ruling it out. The manufacturer’s intended use includes recommendations for use in patients older than 2 years, presenting within 3 days of an acute onset fever (exhibited or reported), and within 7 days of new onset respiratory symptoms consistent with a community-acquired upper respiratory infection (11).

We undertook a systematic review and individual patient data (IPD) meta-analysis to evaluate the diagnostic characteristics of FebriDx (MxA component) compared to contemporaneous reverse transcriptase polymerase chain reaction (RT-PCR) testing for identifying patients with COVID-19 as part of the COVID-19 National Diagnostic Research and Evaluation (CONDOR) test evaluation programme (12). We did not limit this analysis to patients presenting within a certain number of days since the onset of symptoms, in order to inform and identify all potential use cases within the pandemic.

## Method

We performed a systematic review and IPD meta-analysis of diagnostic test accuracy (DTA) studies, and followed the Transparent Reporting of Systematic Reviews and Meta-analyses (PRISMA) extension for DTA (PRISMA-DTA) checklist (13) and PRISMA-DTA for abstracts (14) checklist (supplementary material 1). We also followed the PRISMA-IPD checklist (PRISMA-IPD) (15) (supplementary material 2). This study was conducted at pace as part of the CONDOR test evaluation programme (12), and as a result, no protocol was developed, and the study was not registered.

### Literature search

The inclusion criteria for studies were: [I1] published or un-published (i.e. preprint) DTA studies; [I2] the FebriDx LFD used as the index test; [I3] RT-PCR used as the reference test; and [I4] an adult population suspected of COVID-19 regardless of the time since symptom onset. The exclusion criteria for studies were: [E1] studies that were not a DTA study. Inclusion criteria I2, I3, and I4 were applied at the individual level, where eligible participants could be included and ineligible participants excluded from a study that included a wider population than specified by the criteria.

A number of databases were electronically searched on the 13^th^ of January 2021 by one author (JS), including MEDLINE, EMBASE, BioRxiv and MedRxiv via a Living Systematic Review on SARS-CoV-2 (16), PubMed (17), Google Scholar (18), LitCovid (19), ClinicalTrials.gov (20), and the Living OVerview of Evidence (LOVE) platform (21). Search terms included “FebriDx” AND “COVID-19”, amongst others (see supplementary material 3 for the full search strategy). A date restriction to articles published from 2019 onwards was applied. No location or language restrictions were applied, and all of the databases included pre-prints. No subsequent contact was made with the included study authors for the purpose of detecting additional studies not found by the initial search.

The abstracts of the search results were accessed and screened against the inclusion and exclusion criteria by two authors independently (SGU and KG). The two authors discussed, compared, and combined their findings, and if there was disagreement, adjudication was provided by a third author (AJA). If there was insufficient information in the abstract to exclude the study based on the inclusion and exclusion criteria, then a conservative approach of accessing the full text to perform the screening was taken to mitigate the risk of erroneously excluding relevant studies.

### Individual patient data

The Chief Investigators of the studies that passed eligibility screening against the inclusion and exclusion criteria were approached via email to provide anonymised IPD. If IPD were not available, or not provided, then the study was excluded from further analysis. The minimum data set that was requested is outlined in supplementary material 4. Any queries relating to the study or provided data were communicated and resolved via email.

### Risk of bias and applicability assessments

Risk of bias (RoB) and applicability assessments were performed on the included studies using the QUADAS-2 tool (22) for the quality assessment of diagnostic accuracy studies by two authors independently (SGU and KG). The two authors held a discussion to compare and combine their findings, and if there was disagreement, the final decision was adjudicated by a third author (BCL). If it was decided that the information in the manuscript was insufficient to answer the questions definitively within the QUADAS-2 tool (i.e. an ‘unclear’ rating), then the Chief Investigators of the study were contacted via email to clarify relevant details.

### Data processing

Following receipt of the IPD, the data were checked for completeness by quantifying the amount of missing data for each variable, and for consistency between studies by analysing the definition, data type, and scaling of the common variables. FebriDx and RT-PCR results were summarised into 2×2 contingency tables for each study independently. The following assumptions were applied to dichotomise the outcome of FebriDx and RT-PCR into ‘positive’ and ‘negative’ for the outcome of ‘COVID-19’. For FebriDx: a viral result (i.e. MxA positive [≥40 ng/ml], regardless of CRP positivity) was considered positive; a non-viral result (i.e. MxA negative [<40 ng/ml], CRP positive [≥20mg/L]) was considered negative; and a negative result (i.e. MxA negative [<40 ng/ml], CRP negative [<20mg/L]) was considered negative. For RT-PCR: if SARS-CoV-2 was detected the result was considered positive (COVID-19 + ‘other’ were also considered positive if studies used a respiratory panel that also tested for other viruses); and if SARS-CoV-2 was not detected the result was considered negative.

### Data analysis

For the main analysis, a complete case analysis approach was taken, whereby only patients with a completed and valid FebriDx and RT-PCR result pair were included. Missingness in other variables was not considered for this analysis. This was undertaken to maximise the available sample size for the meta-analysis. Patients that had missing or invalid FebriDx or RT-PCR results were therefore excluded from the main analysis.

To determine if the study populations were similar, and to analyse any baseline and outcome imbalance, the distributions of patient characteristics and outcome variables were quantified for each of the included studies independently. Numerical data were summarised using the median and interquartile range, whilst categorical data were summarised using counts and proportions. The reported test yield was calculated as the proportion of all patients tested with FebriDx that had a valid test result (including after retesting, if applicable) (supplementary material 5). Diagnostic accuracy statistics with 95% confidence intervals (95% CIs) were calculated from the 2×2 contingency tables for each study independently (supplementary material 5).

### Meta-analysis

#### Main analysis

A two-step approach was performed for the IPD meta-analysis. The observed true positive, false positive, true negative and false negative frequencies of the included studies were continuity corrected if values of 0 or 1 would result for the sensitivity or false positive rate, then the sensitivities and false positive rates were transformed using the logit transformation. The bivariate model of Reitsma et al. (23), a linear mixed model with random effects of known variances estimated by restricted maximum likelihood (REML), was fitted to the pairs of transformed sensitivities and false positive rates to obtain a pooled estimate of sensitivity and specificity (1-false positive rate) across included studies using the ‘mada’ R package (24). The absolute value of the true variance (heterogeneity, known as ‘tau-squared’), was used to quantify heterogeneity across studies. A summary receiver operating characteristic curve (SROC) from the resulting model was then plotted with a 95% confidence and prediction region using the ‘mada’ R package (24), with area under the curve (AUC) and partial AUC (restricted to observed false positive rates and normalised) calculated.

#### Sub-group analysis

A sub-group analysis for symptom duration was performed to determine whether there were differences in the pooled estimates of sensitivity and specificity between patients with different symptom durations. Patients with missing symptom duration were excluded. A meta-regression was performed using the same modelling approach as detailed above for the main analysis, but including the covariate symptom duration, with patients dichotomised into two groups (0 to 7 days; >7 days). A sub-group analysis for age was also performed to determine whether there were differences in the pooled estimates of sensitivity and specificity between patients of different ages. Patients with missing age were excluded. A meta-regression was performed using the same modelling approach as detailed above for the main analysis, but including the covariate age, with patients dichotomised into two groups (16 to pooled median years; >pooled median years). Tau-squared was used to quantify heterogeneity across studies. All data processing and analysis was performed in the statistical programming language R (25), using the RStudio (Version 1.2.1335) integrated development environment.

## Results

Sixty-four studies were identified from the literature search, with fifty-four studies excluded following deduplication. Ten studies were screened for eligibility against the inclusion and exclusion criteria, with seven studies excluded, leaving three studies for potential inclusion (supplementary material 6). All three studies were from the UK: a study from Southampton (Chief Investigator = TWC) (6), a study from Kettering (Chief Investigator = RVR) (10), and a study from London (Chief Investigator = LJ) (26). The Chief Investigators of all three studies were able to provide IPD. In addition to receiving the requested IPD, an additional, unpublished dataset was made available to us from the Southampton study using the same protocol as their initial publication (6). For the London study, IPD were only received for the sub-group of patients with an intermediate pre-test probability of COVID-19 which compared the diagnostic accuracy of FebriDx to RT-PCR.

### Study risk of bias and applicability assessment

The results of the RoB and applicability assessment are provided in supplementary material 7. In the ‘Patient selection’ domain, two out of the three studies were considered to have a high risk of bias in relation to the current review, with the Kettering and London studies reporting a large proportion of exclusions. The Kettering study excluded one third of patients (n = 25/75) for having symptoms longer than 7 days (10). Despite this following FebriDx’s instructions for use (11), it is unlikely to reflect clinical use and is a potential source of bias in the current review. The London study excluded approximately 10% of patients (n = 145/1225) due to an unclear reason as to why they weren’t tested with FebriDx, which is a potential source of bias in the current review (26). All three studies were considered to have a low risk of bias across the ‘Index test’ and ‘Reference test’ domains. Due to the aforementioned exclusions in the Kettering and London studies, both were also considered to have a high risk of bias in the ‘Flow and timing’ domain. All studies were considered to be a low concern regarding applicability across all domains (‘Patient selection’, ‘Index test’, and ‘Reference test’) (supplementary material 7).

### Study eligibility criteria, index test, reference standard, and patient flow

Table 1 presents a summary of the eligibility criteria, index test, and reference standard used within the included studies.

**Table 1.**
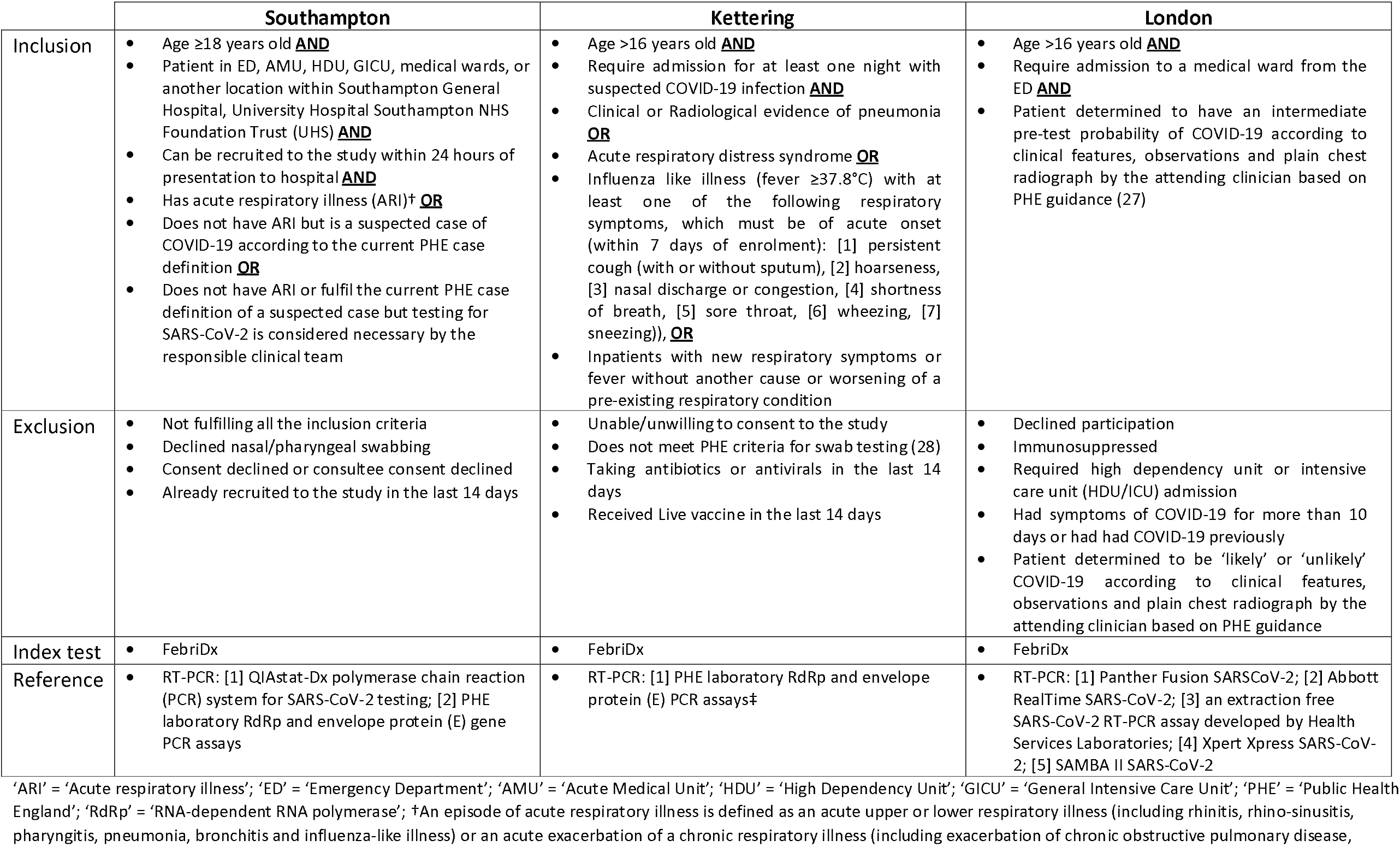
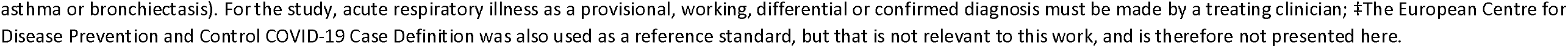
The eligibility criteria, index test, and reference standard of the included studies.

|  | Southampton | Kettering | London |
| --- | --- | --- | --- |
| Inclusion | Age ≥18 years old **AND**<br>Patient in ED, AMU, HDU, GICU, medical wards, or another location within Southampton General Hospital, University Hospital Southampton NHS Foundation Trust (UHS) **AND**<br>Can be recruited to the study within 24 hours of presentation to hospital **AND**<br>Has acute respiratory illness (ARI)† **OR**<br>Does not have ARI but is a suspected case of COVID-19 according to the current PHE case definition **OR**<br>Does not have ARI or fulfil the current PHE case definition of a suspected case but testing for SARS-CoV-2 is considered necessary by the responsible clinical team | Age >16 years old **AND**<br>Require admission for at least one night with suspected COVID-19 infection **AND**<br>Clinical or Radiological evidence of pneumonia **OR**<br>Acute respiratory distress syndrome **OR**<br>Influenza like illness (fever ≥37.8°C) with at least one of the following respiratory symptoms, which must be of acute onset (within 7 days of enrolment): [1] persistent cough (with or without sputum), [2] hoarseness, [3] nasal discharge or congestion, [4] shortness of breath, [5] sore throat, [6] wheezing, [7] sneezing), **OR**<br>Inpatients with new respiratory symptoms or fever without another cause or worsening of a pre-existing respiratory condition | Age >16 years old **AND**<br>Require admission to a medical ward from the ED **AND**<br>Patient determined to have an intermediate pre-test probability of COVID-19 according to clinical features, observations and plain chest radiograph by the attending clinician based on PHE guidance (27) |
| Exclusion | Not fulfilling all the inclusion criteria<br>Declined nasal/pharyngeal swabbing<br>Consent declined or consultee consent declined<br>Already recruited to the study in the last 14 days | Unable/unwilling to consent to the study<br>Does not meet PHE criteria for swab testing (28)<br>Taking antibiotics or antivirals in the last 14 days<br>Received Live vaccine in the last 14 days | Declined participation<br>Immunosuppressed<br>Required high dependency unit or intensive care unit (HDU/ICU) admission<br>Had symptoms of COVID-19 for more than 10 days or had had COVID-19 previously<br>Patient determined to be 'likely' or 'unlikely' COVID-19 according to clinical features, observations and plain chest radiograph by the attending clinician based on PHE guidance |
| Index test | FebriDx | FebriDx | FebriDx |
| Reference | RT-PCR: [1] QIAstat-Dx polymerase chain reaction (PCR) system for SARS-CoV-2 testing; [2] PHE laboratory RdRp and envelope protein (E) gene PCR assays | RT-PCR: [1] PHE laboratory RdRp and envelope protein (E) PCR assays‡ | RT-PCR: [1] Panther Fusion SARSCoV-2; [2] Abbott RealTime SARS-CoV-2; [3] an extraction free SARS-CoV-2 RT-PCR assay developed by Health Services Laboratories; [4] Xpert Xpress SARS-CoV-2; [5] SAMBA II SARS-CoV-2 |

‘ARI’ = ‘Acute respiratory illness’; ‘ED’ = ‘Emergency Department’; ‘AMU’ = ‘Acute Medical Unit’; ‘HDU’ = ‘High Dependency Unit’; ‘GICU’ = ‘General Intensive Care Unit’; ‘PHE’ = ‘Public Health England’; ‘RdRp’ = ‘RNA-dependent RNA polymerase’; †An episode of acute respiratory illness is defined as an acute upper or lower respiratory illness (including rhinitis, rhino-sinusitis, pharyngitis, pneumonia, bronchitis and influenza-like illness) or an acute exacerbation of a chronic respiratory illness (including exacerbation of chronic obstructive pulmonary disease,
asthma or bronchiectasis). For the study, acute respiratory illness as a provisional, working, differential or confirmed diagnosis must be made by a treating clinician; ‡The European Centre for Disease Prevention and Control COVID-19 Case Definition was also used as a reference standard, but that is not relevant to this work, and is therefore not presented here.

#### Eligibility criteria

In the Southampton study, consecutive patients were approached for participation at Southampton General Hospital between the 20^th^ March 2020 and 29^th^ April 2020 (6). In the Kettering study, consecutive patients were approached for participation at Kettering General Hospital between 16^th^ of March and 7^th^ of April 2020 (10). In the London study, patients were included if they required admission to a medical ward from the Emergency Department (ED) at Northwick Park Hospital between 10^th^ of August 2020 and 4^th^ of November 2020 (26). When combined, all studies recruited patients receiving hospital care over 16 years of age with suspected COVID-19 between the 16^th^ of March and 4^th^ November 2020, however the Southampton study also allowed patients to be recruited who did not have an acute respiratory illness (ARI) or did not meet the PHE definition of a suspected case, but where testing was considered necessary by the clinical team (6).

#### Index test

In all studies, the fingerstick blood samples for FebriDx were taken at the same or similar time as the nasopharyngeal swab for RT-PCR. In addition, the readers of the FebriDx test lines were blinded to the RT-PCR results, and vice versa, in all studies. In the Southampton study, FebriDx was performed by research staff and the result was read independently by two study investigators and disagreements were further adjudicated by a third study investigator (6). In the Kettering study, FebriDx was performed and read by one study physician, however if the result was inconclusive or negative, this was further adjudicated by two study physicians (10). In the London study, FebriDx was performed and read by ED Healthcare Assistants following training (26).

#### Reference standard

The Southampton study used the QIAstat-Dx RT-PCR system (Qiagen, Hilden, Germany) for analysing nasopharyngeal swabs, which gave a binary readout of positive or negative for the detection of targets including SARS-CoV-2 (6), in addition to Public Health England (PHE) laboratory RNA-dependent RNA polymerase (RdRp) and envelope protein (E) RT-PCR testing for SARS-CoV-2 (6). However, only results from the QIAstat-Dx RT-PCR system were available for the additional unpublished data from the Southampton study following their publication (6), so the results from the QIAstat-Dx RT-PCR system were used for all patients from the Southampton study to maintain within-study consistency for the purposes of the current review. The Kettering study used PHE laboratory RdRp and envelope protein (E) RT-PCR testing for SARS-CoV-2 to analyse nasopharyngeal swabs (10). The London study used the Panther Fusion RT-PCR system (Hologic Inc, CA, USA), the Abbott RealTime system (Abbott Park, IL, USA), an extraction free RT-PCR assay developed by Health Services Laboratories in the UK, the Xpert Xpress RT-PCR system (Cepheid, CA, USA), and the SAMBA II RT-PCR system (Diagnostics for Real World, CA, USA) for analysing nasopharyngeal swabs for the detection of SARS-CoV-2.

#### Patient flow

The flow of patients in the Southampton, Kettering, and London studies for the IPD included in the current review are summarised in supplementary material 8, 9, and 10, respectively. In the Southampton study, 500 patients were approached for testing with FebriDx, with 22 excluded (4.4%) as it was deemed inappropriate by the clinical team, or where the patient/carer declined participation in the study. Out of the 478 patients tested with FebriDx, 19 tests were initially invalid (4%). FebriDx could not be repeated in 3 of the 19 initially invalid tested patients (15.8%). Out of the 16 initially invalid tested patients that were retested, 1 was invalid (6.3%), and was subsequently retested again where the patient then received a valid test result upon a second retest. Considering all 20 invalid tests, 16 were due to blood clotting in the collection tube (80%), whilst 4 were due to there being no CRP line [<20mg/L] on the FebriDx device (20%). 475 patients remained for analysis, resulting in a reported test yield of 99.4% (supplementary material 8). The Kettering study approached 75 patients for testing with FebriDx, where 26 were excluded (34.7%), with 25 due to symptoms being longer than 7 days, and 1 due to being immunosuppressed. Out of the 49 patients tested with FebriDx, 1 test was initially invalid (2%) due to the inability to obtain enough blood. FebriDx could not be repeated in this patient as they were elderly, frail, and clinically unstable at the time of testing, and 48 patients remained for analysis, resulting in a reported test yield of 98% (supplementary material 9). The London study included 3433 adult medical admissions triaged through the ED who had RT-PCR test results. Of these patients, 1225 were deemed to have an intermediate pre-test probability of COVID-19, where 267 were excluded (21.8%) due to FebriDx not being performed, with 145 due to an unclear reason why FebriDx was not performed, 60 due to being immunosuppressed, 27 due to needing a higher level of care, 20 due to symptoms being longer than 10 days, 13 due to having previous COVID-19, 1 due to being unable to bleed, and 1 due to refusing FebriDx. 958 patients were tested with FebriDx and remained for analysis, resulting in a reported test yield of 100% (supplementary material 10).

### Study patient characteristics and outcomes

The patient characteristics and outcomes for the included studies are summarised in Table 2.

**Table 2.**
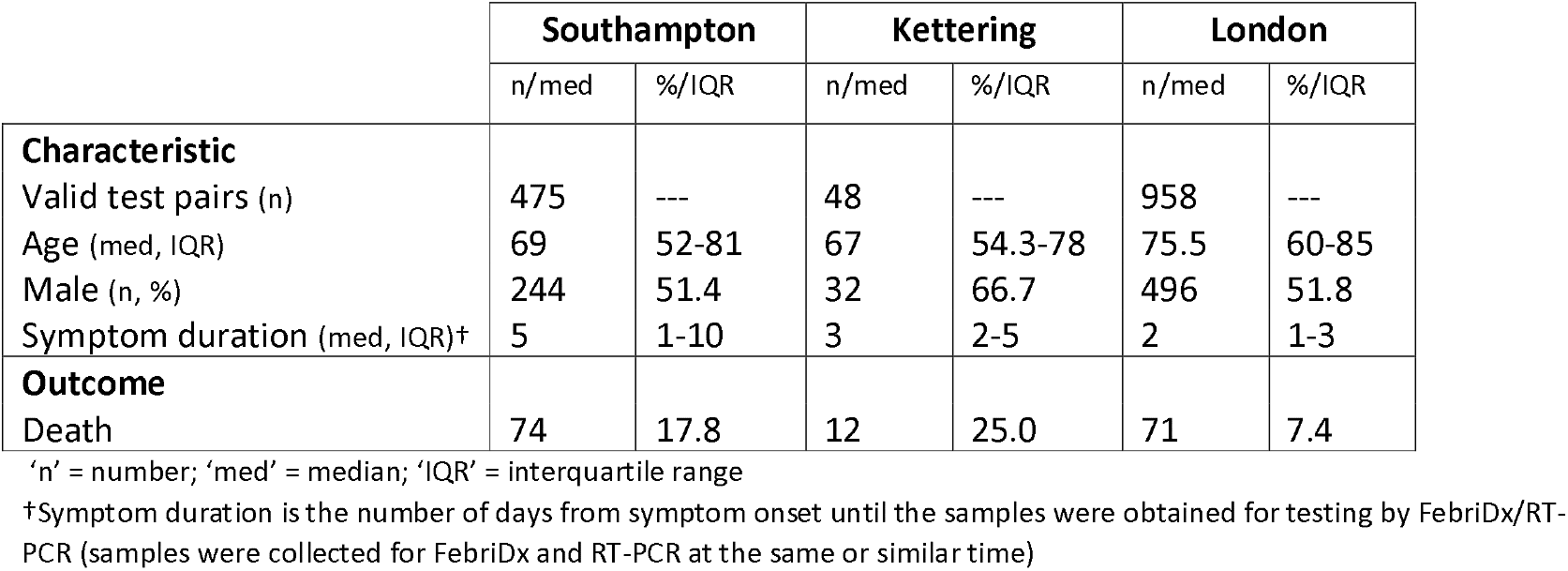
Patient characteristics and outcomes in the included studies.

|  | Southampton |  | Kettering |  | London |  |
| --- | --- | --- | --- | --- | --- | --- |
|  | n/med | %/IQR | n/med | %/IQR | n/med | %/IQR |
| **Characteristic** |  |  |  |  |  |  |
| Valid test pairs (n) | 475 | --- | 48 | --- | 958 | --- |
| Age (med, IQR) | 69 | 52-81 | 67 | 54.3-78 | 75.5 | 60-85 |
| Male (n, %) | 244 | 51.4 | 32 | 66.7 | 496 | 51.8 |
| Symptom duration (med, IQR)† | 5 | 1-10 | 3 | 2-5 | 2 | 1-3 |
| **Outcome** |  |  |  |  |  |  |
| Death | 74 | 17.8 | 12 | 25.0 | 71 | 7.4 |

‘n’ = number; ‘med’ = median; ‘IQR’ = interquartile range
†Symptom duration is the number of days from symptom onset until the samples were obtained for testing by FebriDx/RT-PCR (samples were collected for FebriDx and RT-PCR at the same or similar time)

Patients in the Southampton and Kettering studies had a similar age distribution, with a median of 69 and 67 years, respectively. Patients in the London study were older with a median of 75.5 years (Table 2). The Southampton and London studies had a similar sex distribution, where 51.4 and 51.8% of patient were male, respectively. The Kettering study had a higher proportion of male patients at 66.7%. Patients in the Kettering and London studies had a similar symptom duration distribution, with a median of 3 and 2 days, respectively; whilst patients in the Southampton study had a longer symptom duration with a median of 5 days. The Southampton and Kettering studies had a similar mortality rate of 17.8 and 25%, respectively. The London study had a lower mortality rate of 7.4%. The Southampton study reported death at 30 days following admission, whereas the Kettering and London studies reported death at the end of the index admission.

### Diagnostic accuracy

The Southampton, Kettering, and London studies had a reported FebriDx test yield of 99.4%, 98%, and 100%, respectively. These valid tests are presented in 2×2 contingency tables for each of the studies in Figure 1, with the resulting prevalence and diagnostic accuracy statistics presented in Table 3.

**Table 3.**
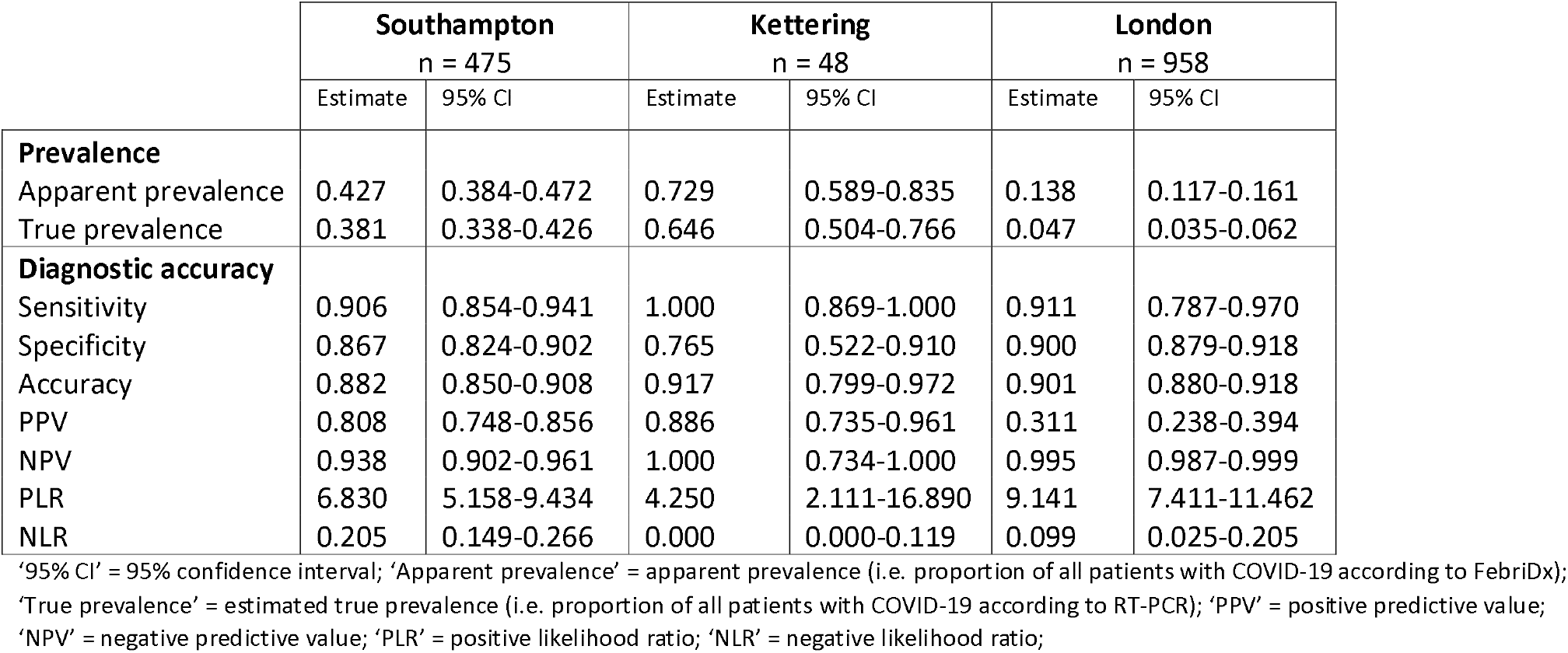
Prevalence and diagnostic accuracy statistics for the included studies.

|  | Southampton<br>n = 475 |  | Kettering<br>n = 48 |  | London<br>n = 958 |  |
| --- | --- | --- | --- | --- | --- | --- |
|  | Estimate | 95% CI | Estimate | 95% CI | Estimate | 95% CI |
| **Prevalence** |  |  |  |  |  |  |
| Apparent prevalence | 0.427 | 0.384-0.472 | 0.729 | 0.589-0.835 | 0.138 | 0.117-0.161 |
| True prevalence | 0.381 | 0.338-0.426 | 0.646 | 0.504-0.766 | 0.047 | 0.035-0.062 |
| **Diagnostic accuracy** |  |  |  |  |  |  |
| Sensitivity | 0.906 | 0.854-0.941 | 1.000 | 0.869-1.000 | 0.911 | 0.787-0.970 |
| Specificity | 0.867 | 0.824-0.902 | 0.765 | 0.522-0.910 | 0.900 | 0.879-0.918 |
| Accuracy | 0.882 | 0.850-0.908 | 0.917 | 0.799-0.972 | 0.901 | 0.880-0.918 |
| PPV | 0.808 | 0.748-0.856 | 0.886 | 0.735-0.961 | 0.311 | 0.238-0.394 |
| NPV | 0.938 | 0.902-0.961 | 1.000 | 0.734-1.000 | 0.995 | 0.987-0.999 |
| PLR | 6.830 | 5.158-9.434 | 4.250 | 2.111-16.890 | 9.141 | 7.411-11.462 |
| NLR | 0.205 | 0.149-0.266 | 0.000 | 0.000-0.119 | 0.099 | 0.025-0.205 |

'95% CI' = 95% confidence interval; 'Apparent prevalence' = apparent prevalence (i.e. proportion of all patients with COVID-19 according to FebriDx); 'True prevalence' = estimated true prevalence (i.e. proportion of all patients with COVID-19 according to RT-PCR); 'PPV' = positive predictive value; 'NPV' = negative predictive value; 'PLR' = positive likelihood ratio; 'NLR' = negative likelihood ratio;

**Figure 1.**
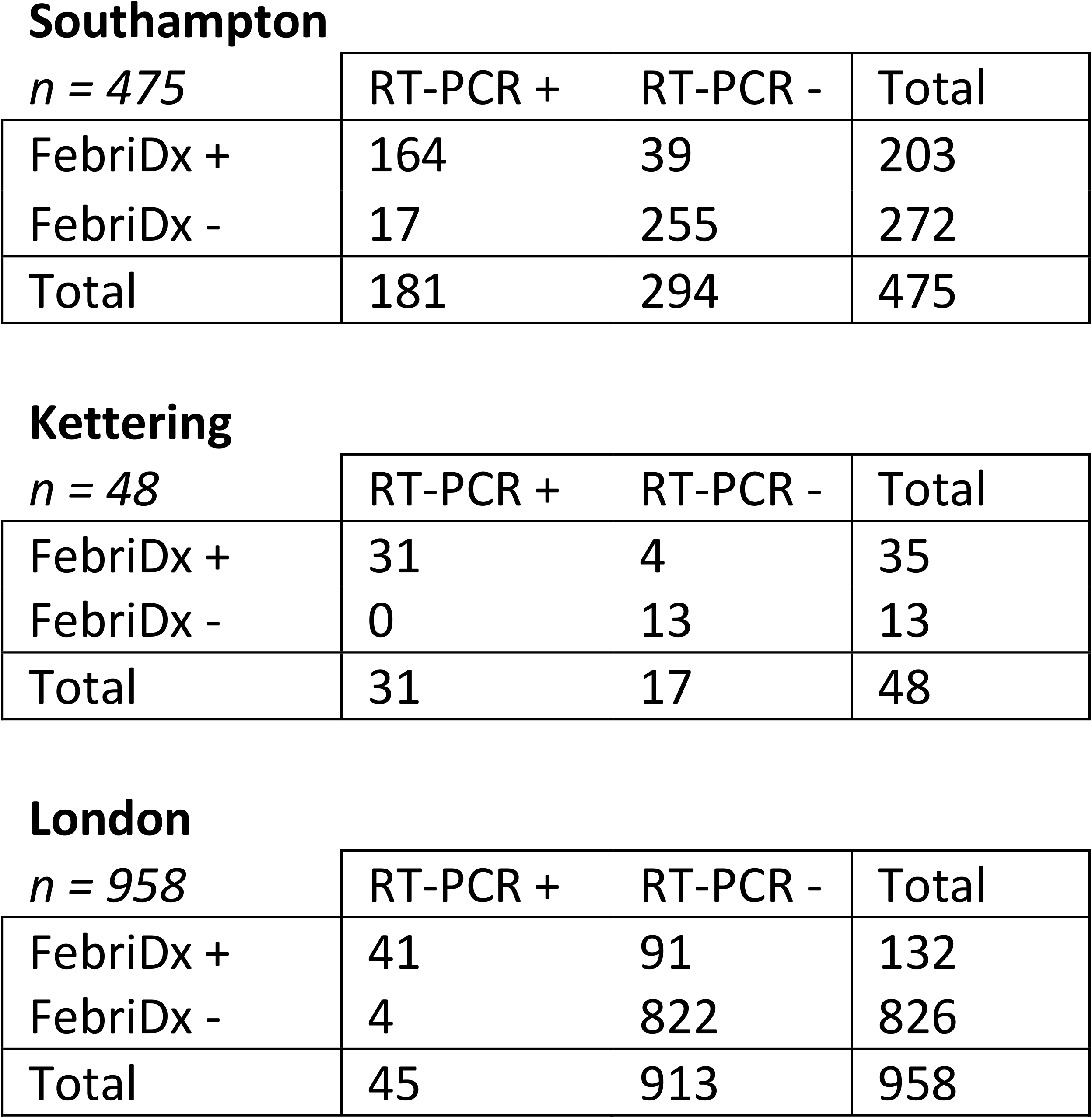
2×2 contingency tables from the three included studies showing the diagnostic accuracy results of FebriDx versus RT-PCR for COVID-19

The estimated true prevalence of COVID-19, i.e. the proportion of all patients with COVID-19 according to RT-PCR, varied amongst studies (Table 3). The Southampton study had an estimated true prevalence of 0.381 (95% CI: 0.338-0.426), the Kettering study had a higher estimated true prevalence of 0.646 (95% CI: 0.504-0.766), and the London study had a lower estimated true prevalence of 0.047 (95% CI: 0.035-0.062). The Southampton and London studies were found to have a similar sensitivity, with a point estimate of 0.906 (95% CI: 0.854-0.941) and 0.911 (95% CI: 0.787-0.970), respectively. The Kettering study was found to have a higher sensitivity point estimate of 1.000 (95% CI: 0.869-1.000). The Southampton and London studies were also found to have a similar specificity, with a point estimate of 0.867 (95% CI: 0.824-0.902) and 0.900 (95% CI: 0.879-0.918), respectively. The Kettering study was found to have a lower specificity point estimate of 0.765 (95% CI: 0.522-0.910) (Table 3).

### Meta-analysis

#### Main analysis

The bivariate model included three studies, and had two fixed and three random effects parameters. The two fixed effect coefficients were: transformed sensitivity (intercept = 2.3214) and transformed false positive rate (intercept = −1.8852). The three random effects parameters were: log likelihood (10.9274), Akaike information criterion (AIC) (−11.8548), and Bayesian information criterion (BIC) (−12.8960). The model produced a pooled sensitivity estimate of 0.911 (95% CI: 0.855-0.946) and pooled specificity estimate of 0.868 (95% CI: 0.802-0.915). The variance (between study standard deviation, known as ‘tau’) was estimated to be 0.256 for sensitivity and 0.356 for specificity. The absolute value of the true variance (heterogeneity, known as ‘tau-squared’), was estimated to be lower for sensitivity (0.066) than specificity (0.127). A SROC of the model is presented in Figure 2. The model produced an AUC of 0.946, and partial AUC of 0.914.

**Figure 2.**
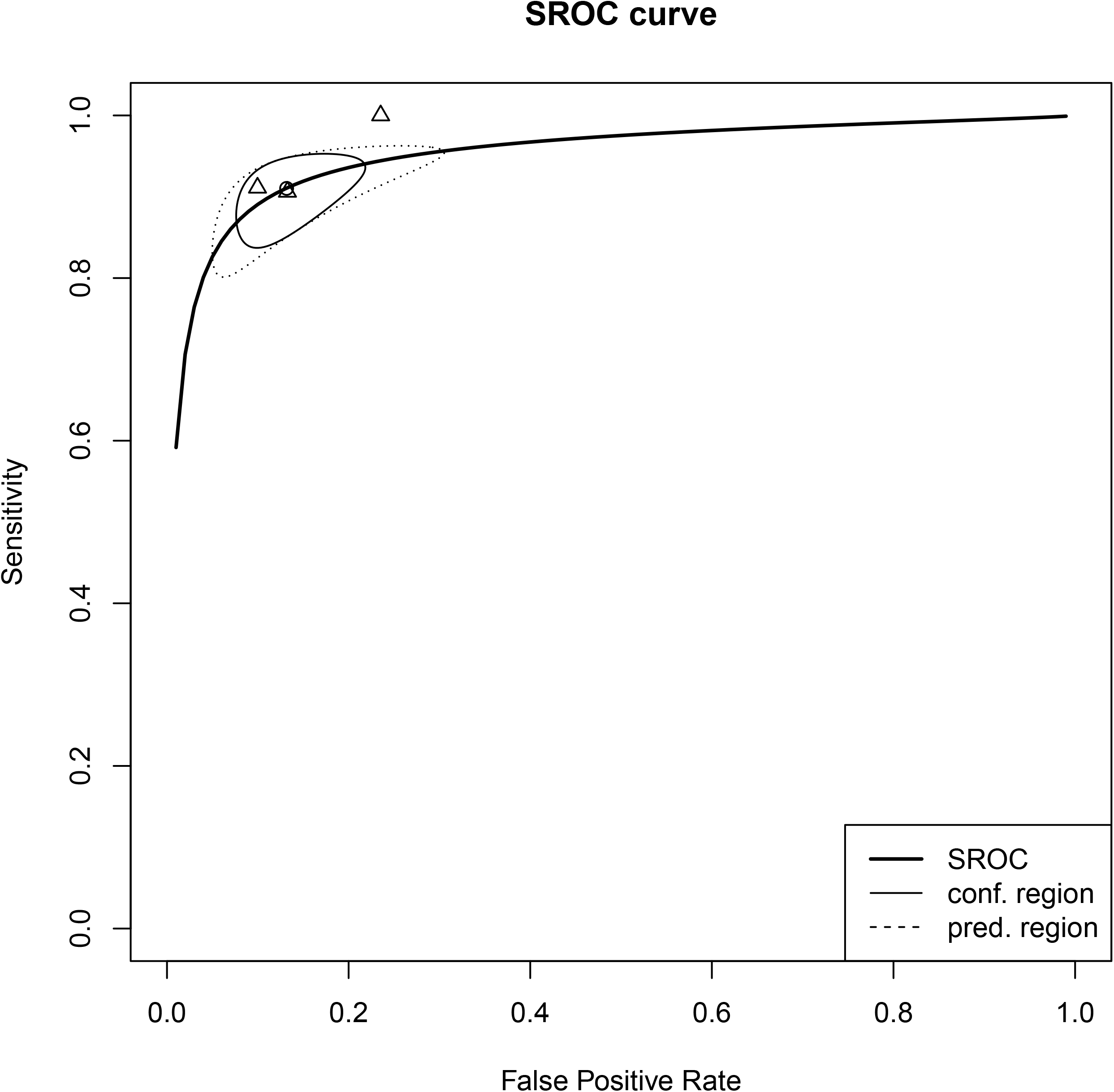
Summary receiver operating characteristic (SROC) curve of the bivariate model of Reitsma et al. (26), a linear mixed model with random effects, which was fitted to obtain a pooled estimate of sensitivity and specificity from the three included studies (identified as the circle on the plot: sensitivity = 0.911 (95% CI: 0.855-0.946), specificity = 0.868 (95% CI: 0.802-0.915)). A 95% confidence and prediction region around this pooled estimate is outlined by a straight and dashed line, respectively. The individual study estimates are identified by triangles.

#### Sub-group analysis

The bivariate meta-regression model for symptom duration included three studies (although the Kettering study did not have any patients with a symptom duration of >7 days), and had four fixed and three random effects parameters. The four fixed effect coefficients were: transformed sensitivity (intercept = 1.9278; group 0 to 7 days = 0.4572) and transformed false positive rate (intercept = −1.9562; group 0 to 7 days = 0.0960). The three random effects parameters were: log likelihood (17.2890), AIC (−20.5780), and BIC (−18.4599). There were no statistically significant differences between the groups of 0 to 7 days and >7 days in estimated pooled sensitivity (p = 0.473) or estimated pooled specificity (p = 0.853). Tau was estimated to be 0.424 for sensitivity and 0.442 for specificity. Tau-squared was estimated to be similar for sensitivity (0.180) and specificity (0.195).

The bivariate meta-regression model for age included three studies, and had four fixed and three random effects parameters. The four fixed effect coefficients were: transformed sensitivity (intercept = 2.2644; group 16 to 73 years = −0.0327) and transformed false positive rate (intercept = −2.0508; group 0 to7 days = 0.2725). The three random effects parameters were: log likelihood (19.5211), AIC (−25.0422), and BIC (−21.6478). There were no statistically significant differences between the groups of 16 to 73 years of age and >73 years of age in estimated pooled sensitivity (p = 0.946) or estimated pooled specificity (p = 0.486). Tau was estimated to be 0.130 for sensitivity and 0.363 for specificity. Tau-squared was estimated to be lower for sensitivity (0.017) than specificity (0.132).

## Discussion

In this systematic review and IPD meta-analysis of DTA studies, we found that the FebriDx LFD had a pooled sensitivity estimate of 0.911 (95% CI: 0.855-0.946) and specificity of 0.868 (95% CI: 0.802-0.915) for identifying COVID-19 using the MxA biomarker compared to RT-PCR for the SARS-CoV-2 virus. Three studies were included, with a total of 1481 patients recruited within acute hospitals in the UK. There was no evidence of age and symptom duration having an impact on the diagnostic accuracy of FebriDx, with no statistically significant differences between the two age sub-groups of 16 to 73 years of age and >73 years of age, and the two symptom duration sub-groups of 0 to 7 days and >7 days. However, only two out of the three studies included patients with a symptom duration of >7 days, so this finding is based on limited data.

The RoB assessment identified that two out of the three studies were at high risk of bias in the ‘Patient selection’ and ‘Flow and timing’ domains due to reporting a large proportion of exclusions, with the Kettering study excluding one third of patients due to having symptoms longer than 7 days, and the London study excluding approximately 10% of patients due to an unclear reason why they weren’t tested with FebriDx. This, coupled with the small evidence base, where the estimates of the variances of the random effects will be subject to a high level of uncertainty, suggests that the findings should be interpreted with caution despite observing low to moderate heterogeneity between studies in the meta-analysis outcomes.

The patients included in the three studies were from a number of acute and inpatient hospital settings with varying COVID-19 prevalence, and such findings must be extrapolated with caution to more specific patient groups and settings both within and outside of hospital. This may be particularly true for community settings on the basis that patients with mild/moderate COVID-19 not requiring hospital admission may be less likely to have a measurable Type I or Type III interferon response because of their disease severity. Further context-specific evaluation would be required in order for FebriDx to be used in other patient groups where performance has not yet been demonstrated; such as children, immunocompromised and cancer patients, those who are asymptomatic, and care home residents. Taking care homes as an example, the mean age of residents is 85 years (29), and all care home residents are significantly affected by frailty. The high prevalence of immunosenescence in this group is such that MxA and CRP results might be significantly attenuated (30). It is clear that the current hospital data cannot be extrapolated to such a group and further context-specific evaluation would be required.

In the context of older, frailer, community dwelling populations where delirium is a common and sensitive, but non-specific presentation of COVID-19 (31), the ability to rule out viral and bacterial infections as the cause of delirium during an outbreak may be even more important than the ability to detect them. Further work is therefore required to look at the sensitivity and negative predictive value of FebriDx in populations where such information may be of use. Specific treatments (neuraminidase inhibitors) are available and recommended for use during influenza outbreaks, and an increasing body of evidence supports different interventions (remdesivir, dexamethasone, and budesonide) for use in cases of COVID-19 (32-34).

Although not a direct test for the presence of SARS-CoV-2 infection, the performance measures reported in our analysis are comparable with results from other studies of FebriDx in detecting the presence of viral respiratory infections (7). As a raised MxA level is not diagnostic of SARS-CoV-2 due to its non-specific response to a number of respiratory infections, the optimal use case of FebriDx is unlikely to be for ‘ruling in’ COVID-19. The simplicity of a fingerstick blood test with a 10-minute turnaround time could enable rapid ‘rule out’ of COVID-19 in patients who have low concentrations of MxA. In a hospital setting, those patients could be potentially sent to non-COVID areas of a hospital, supporting the appropriate use of isolation facilities that could result in significant savings to hospitals which have delays in time to RT-PCR result (35). If FebriDx was used to cohort patients to wards incorrectly, then the unspecific nature of the result may lead to the exposure of patients to potentially serious co-infection. Recent evidence has suggested that the risk of death from co-infection of SARS-CoV-2 and influenza was nearly double that of SARS-CoV-2 alone, 43.1% vs 26.9% (36).

### Implications for practice

Whilst diagnostic performance measures were high, our findings are limited by the uncertain generalisability to longer durations of symptoms (more than a week), to different settings, and to different phases of the pandemic as prevalence rates will vary further. The utility of FebriDx may be limited to its ability to rule out acute COVID-19 infection because a positive result does not specify which viral respiratory pathogen is present and, due to this, the utility may change seasonally because of increasing prevalence of other respiratory viruses. However, a sensitivity of 0.911 will lead to a false negative result in almost one in ten patients with COVID-19. This is an important consideration, notably in settings with a high prevalence of disease. Testing protocols would need to be developed carefully to ensure the correct use of the test, likely as an initial triage test in conjunction with RT-PCR as a confirmatory test.

If further evidence confirms the diagnostic characteristics of FebriDx in a hospital setting then it may have the greatest utility when deployed in a triaging capacity. For example, enabling the allocation of patients to wards based on the likely risk of SARS-CoV-2 whilst confirmatory RT-PCR testing is sought. This should, however, be used with caution due to the potential increased risk that co-infection poses to patients with SARS-CoV-2. The use of FebriDx should be carefully considered within the context of both clinical pathway needs and the patient pathways that it may influence.

### Limitations

The reference standard of RT-PCR on nose and throat swab samples is imperfect, and while commonly used as a reference test, it is not a gold standard (37). RT-PCR has shown limited diagnostic performance characteristics, particularly with the production of false negative results in patients presenting in an emergency with suspected COVID-19 (38-40). An imperfect reference standard in this case, which most likely produced false negative results, would be likely to produce an underestimate of both sensitivity and specificity. If additional clinical and diagnostic data were available for the included studies, this analysis would have benefitted from the use of a composite reference standard (41) or latent class analyses with instrumental variables to minimise the probability of such error or bias (42, 43). Although all studies used RT-PCR as the reference standard, different RT-PCR tests were used within and between studies. The accuracy of the different RT-PCR tests used will vary between methods, and as these methods were further developed during the pandemic, this leads to an inconsistent reference standard.

In our analysis, we only considered the MxA component of FebriDx. It is unclear whether our interpretation of a raised MxA level irrespective of the CRP level is correct as a proxy indicator of COVID-19 and, vice versa, whether our interpretation of a normal MxA level irrespective of the CRP level is correct as a proxy indicator of not having COVID-19. In practice, clinicians may be faced with a challenge in the interpretation of FebriDx results indicating a raised CRP and normal MxA in the context of symptoms consistent with COVID-19.

## Conclusion

Our systematic review and IPD meta-analysis of three studies found that the FebriDx LFD had a high diagnostic accuracy for COVID-19 in varied UK hospital settings, however, the pooled estimates of sensitivity and specificity should be interpreted with caution due to the small number of studies included, risk of bias, and inconsistent reference standards used. FebriDx is likely to be best suited as an initial triage test in conjunction with RT-PCR as a confirmatory test to rule out COVID-19; however, further research is required to confirm these findings before definitive recommendations can be made.

## Data Availability

An anonymised minimum dataset containing enough information to reproduce the diagnostic accuracy statistics is available from the corresponding author on reasonable request. The complete anonymised dataset is not currently available as it is still being used by the study teams to produce further publications.

## List of abbreviations

AMU: Acute Medical Unit
ARI: Acute respiratory illness
CI: Confidence intervals
COVID-19: Coronavirus disease 2019
CRP: C-reactive protein
ED: Emergency Department
GICU: General Intensive Care Unit
HDU: High Dependency Unit
IPD: Individual patient data
IQR: Interquartile range
LFD: Lateral flow device
MxA: Myxovirus resistance protein A
NLR: Negative likelihood ratio
NPV: Negative predictive value
p: P value
PHE: Public Health England
PLR: Positive likelihood ration
POC: Point-of-care
PPV: Positive predictive value
RT-PCR: Reverse transcriptase polymerase chain reaction
RdRp: RNA-dependent RNA polymerase
REML: Restricted maximum likelihood
RoB: Risk of bias
ROC: Receiver operating characteristic
SARS-CoV-2: Severe acute respiratory syndrome coronavirus 2
SOB: Shortness of breath
UK: United Kingdom

## Declarations

### Ethics approval and consent to participate

This work was based on anonymised data from three previous studies; the CoV-19POC study, described by Clark et al. (6), the “Southampton study” [ISRCTN:14966673, date registered: 18/03/2020]; a study described by Karim et al. (10), the “Kettering study”; and a study described by Houston et al. (26), the “London study”. The Southampton study was approved by the South Central - Hampshire A Research Ethics Committee: REC reference 20/SC/0138, on the 16th March 2020. The protocol is available at: https://eprints.soton.ac.uk/439309/1/CoV _ 19POC _ Protocol _ v1.1_eprints.pdf. The Kettering study was approved by the Kettering General Hospital Ethics Committee. The London study was approved by the London North West University Healthcare NHS Trust Research and Development Committee, but due to the study being a retrospective review of routinely collected data, ethical approval was not required. Informed consent was obtained from all patients in the Southampton and Kettering studies, and failure to consent was considered an exclusion criterion in both studies.

### Consent for publication

Informed consent was obtained from all eligible patients.

### Availability of data and materials

Data to reproduce the main and sub-group meta-analyses are provided in the manuscript and supplementary material, respectively.

### Authors’ contributions

RB and AJA devised the project. SGU, BCL and AJA designed the analysis plan. JS designed and executed the literature search. SGU and KG conducted the RoB assessment. SGU conducted the analysis with supervision from BCL and AJA. SGU drafted the initial manuscript. All authors contributed to drafts and revisions of the manuscript.

## Acknowledgements

The authors would like to acknowledge Lumos Diagnostics for sharing information vital to the design of the study, however they had no part in the study design, analysis or of the development of the manuscript.

## Supplementary material

Supplementary material 1: ‘Supplementary material 1.docx’ presents the ‘PRISMA-DTA’ and ‘PRISMA-DTA for abstracts’ checklists.

Supplementary material 2: ‘Supplementary material 2.docx’ presents the ‘PRISMA-IPD’ checklist.

Supplementary material 3 ‘Supplementary material 3.docx’ presents the literature search strategy.

Supplementary material 4: ‘Supplementary material 3.docx’ presents the minimum data set that was requested from the Chief Investigators (CIs) of the included studies.

Supplementary material 5: ‘Supplementary material 4.docx’ presents the methods for calculating the diagnostic accuracy measures.

Supplementary material 6: ‘Supplementary material 5.docx’ presents the PRISMA-IPD flow diagram.

Supplementary material 7: ‘Supplementary material 6.docx’ presents risk of bias (RoB) assessments for the included studies.

Supplementary material 8: ‘Supplementary material 7.docx’ presents the flow of patients in the Southampton study.

Supplementary material 9: ‘Supplementary material 8.docx’ presents the flow of patients in the Kettering study.

Supplementary material 10: ‘Supplementary material 10.docx’ presents the flow of patients in the London study.

Supplementary material 11: ‘Supplementary material 11.docx’ presents the 2×2 contingency tables from the Southampton study, Kettering study, and London study for the symptom duration and age sub-groups.

