## Supplementary material 3 for "FebriDx point-of-care test in patients with suspected COVID-19: a systematic review and individual patient data meta-analysis of diagnostic test accuracy studies"

Literature search strategy.

We conceptualised the searches as: A: [*FebriDx*] AND B: [*COVID-19*]. For element A we used the commercial name of the test. For element B we used search filter for COVID-19 as instructed by Shokraneh (1).

| Database | Search terms | Number of articles |
| --- | --- | --- |
| Living Systematic Review on SARS-CoV-2 | “FebriDx” | 6 |
| PubMed | “FebriDx” AND ((“Betacoronavirus”[Mesh] OR “Coronavirus Infections”[MH] OR “Spike Glycoprotein, COVID-19 Virus”[NM] OR “COVID-19”[NM] OR “Coronavirus”[MH] OR “Severe Acute Respiratory Syndrome Coronavirus 2”[NM] OR “2019nCoV”[ALL] OR “Betacoronavirus”*[ALL] OR “Corona Virus”*[ALL] OR “Coronavirus”*[ALL] OR “Coronovirus”*[ALL] OR “CoV”[ALL] OR “CoV2”[ALL] OR “COVID”[ALL] OR “COVID19”[ALL] OR “COVID-19”[ALL] OR “HCoV-19”[ALL] OR “nCoV”[ALL] OR “SARS CoV 2”[ALL] OR “SARS2”[ALL] OR “SARSCoV”[ALL] OR “SARS-CoV”[ALL] OR “SARS-CoV-2”[ALL] OR “Severe Acute Respiratory Syndrome CoV”*[ALL]) AND ((2019/11/17[EDAT] : 3000[EDAT]) OR (2019/11/17[PDAT] : 3000[PDAT]))) | 3 |
| Google Scholar | “FebriDx” AND “COVID-19” | 22 |
| Google Scholar | “FebriDx” AND allintitle: “COVID-19” OR “COVID19” OR “2019nCoV” OR ”Corona Virus” OR “Coronavirus” OR ”CoV 2” OR “CoV2” OR “COVID” OR “nCoV” OR “SARS2” OR “SARSCoV” OR ”SARS-CoV” since 2019 | 21 |
| LitCovid | “FebriDx” AND “COVID-19” | 3 |
| ClinicalTrials.gov | “FebriDx” AND “COVID-19” | 3 |
| Living OVerview of Evidence (LOVE) | “FebriDx” | 6 |
