## Supplementary material 4 for "FebriDx point-of-care test in patients with suspected COVID-19: a systematic review and individual patient data meta-analysis of diagnostic test accuracy studies"

The minimum data set that was requested from the Chief investigators (CIs) of the included study:

| **Data fields** | **Notes** |
| --- | --- |
| Age | Age in years at time of testing |
| Sex | Male/female |
| Symptom duration | Time tested since the onset of symptoms |
| FebdriDx CRP | Positive/negative; include repeat testing |
| FebdriDx MxA | Positive/negative; include repeat testing |
| FebdriDx control | Positive/negative; include repeat testing |
| FebriDx result | Viral/bacterial/negative; include repeat testing |
| SARS COV-2 PCR result | Positive/negative; include repeat testing |
| Outcome | Discharged home/died/still an inpatient |
