## Supplementary material 5 for "FebriDx point-of-care test in patients with suspected COVID-19: a systematic review and individual patient data meta-analysis of diagnostic test accuracy studies"

The following formula was used to calculate the reported test yield:

**Test yield**

$$Test yield= \frac{n patients with a valid FebriDx test (after retesting, if applicable)}{n patients tested with FebriDx}$$

The following diagnostic accuracy measures were calculated for each of the included studies independently:

- **Apparent prevalence** (the proportion of all patients with COVID-19 according to FebriDx)
- **Estimated true prevalence** (proportion of all patients with COVID-19 according to RT-PCR)
- **Test sensitivity** (proportion of patients with COVID-19, according to RT-PCR, that test positive by FebriDx)
- **Test specificity** (proportion of patients without COVID-19, according to RT-PCR, that test negative by FebriDx)
- **Diagnostic accuracy** (proportion of all FebriDx tests that give a correct result according to RT-PCR)
- **Positive predictive value (PPV)** (probability that patients with a positive FebriDx test have COVID-19 according to RT-PCR)
- **Negative predictive value (NPV)** (probability that patients with a negative FebriDx test do not have the disease according to RT-PCR)
- **Positive likelihood ratio** (probability of a patient who has COVID-19 according to PCR testing positive by FebriDx, divided by the probability of a patient who does not have the disease according to RT-PCR testing positive by FebriDx)
- **Negative likelihood ratio** (probability of a patient who has COVID-19 according to RT-PCR testing negative by FebriDx, divided by the probability of a patient who does not have the disease according to RT-PCR testing negative by FebriDx)

The following formulas were used to calculate the diagnostic accuracy statistics, where ‘TP’ = ‘number of true positives’, ‘FP’ = ‘number of false positives’, ‘TN’ = ‘number of true negatives’, and ‘FN’ = ‘number of false negatives’:

**Apparent prevalence**

$$Apparent prevalence= \frac{TP+FP}{TP+TN+FP+FN}$$

**Estimated true prevalence**

$$Estimated true prevalence= \frac{TP+FN}{TP+TN+FP+FN}$$

**Test sensitivity**

$$Test sensitivity= \frac{TP}{TP+FN}$$

**Test specificity**

$$Test specificity= \frac{TN}{TN+FP}$$

**Diagnostic accuracy**

$$Diagnostic accuracy= \frac{TP+TN}{TP+TN+FP+FN}$$

**Positive predictive value (PPV)**

$$PPV= \frac{TP}{TP+FP}$$

**Negative predictive value (NPV)**

$$NPV= \frac{TN}{TN+FN}$$

Positive and negative likelihood ratios were calculated using the 'diagCI' function of the 'bootLR' R package (1). The Agresti-Coull method was used to compute 95% confidence intervals (CIs) for apparent prevalence, estimated true prevalence, test sensitivity, test specificity, diagnostic accuracy, positive predictive value, and negative predictive value using the 'BinomCI' function of the 'DescTools' R package (2). A Bayesian approach using a bootstrapping method was used to compute 95% CIs for positive and negative likelihood ratios using the 'DiagCI' function of the 'bootLR' package (1) (Marill & Friedman, 2019). All data processing and analysis was performed in the statistical programme language R (3), using the RStudio integrated development environment (4).

3.	R Core Team. R: A language and environment for statistical computing. Vienna, Austria  R Foundation for Statistical  Computing; 2020.

4.	RStudio Team. RStudio: Integrated Development for R B oston, MA, USA: R*Studio, inc.*; 2018 [Available from: http://www.rstudio.com/.
