## Supplementary figures and images for "FebriDx point-of-care test in patients with suspected COVID-19: a systematic review and individual patient data meta-analysis of diagnostic test accuracy studies"

### Supplementary material 6

**Supplementary material 6**

PRISMA-IPD flow diagram.

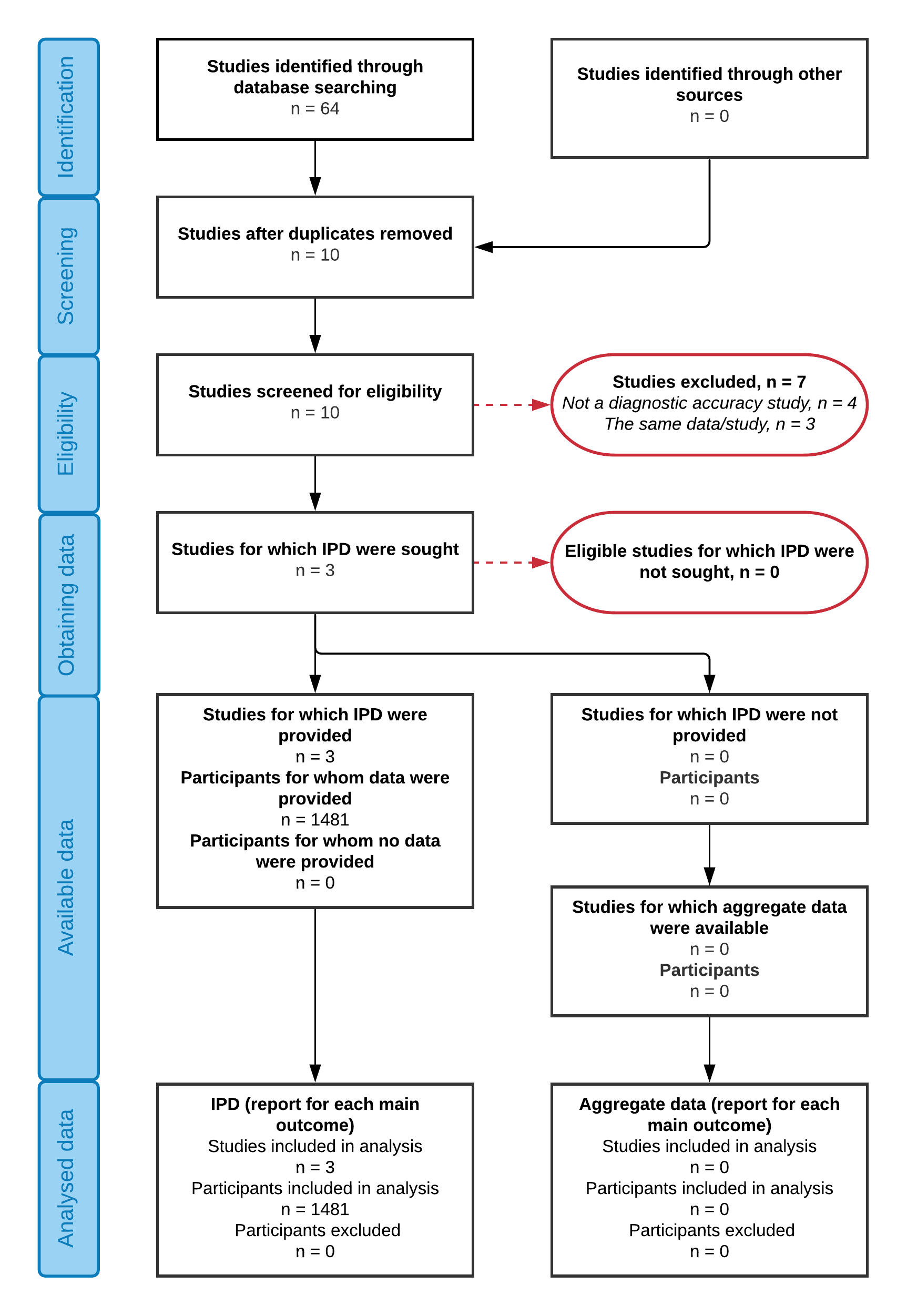

### Supplementary material 9

**Supplementary material 9**

The flow of patients in the Kettering study (Karim et al.):

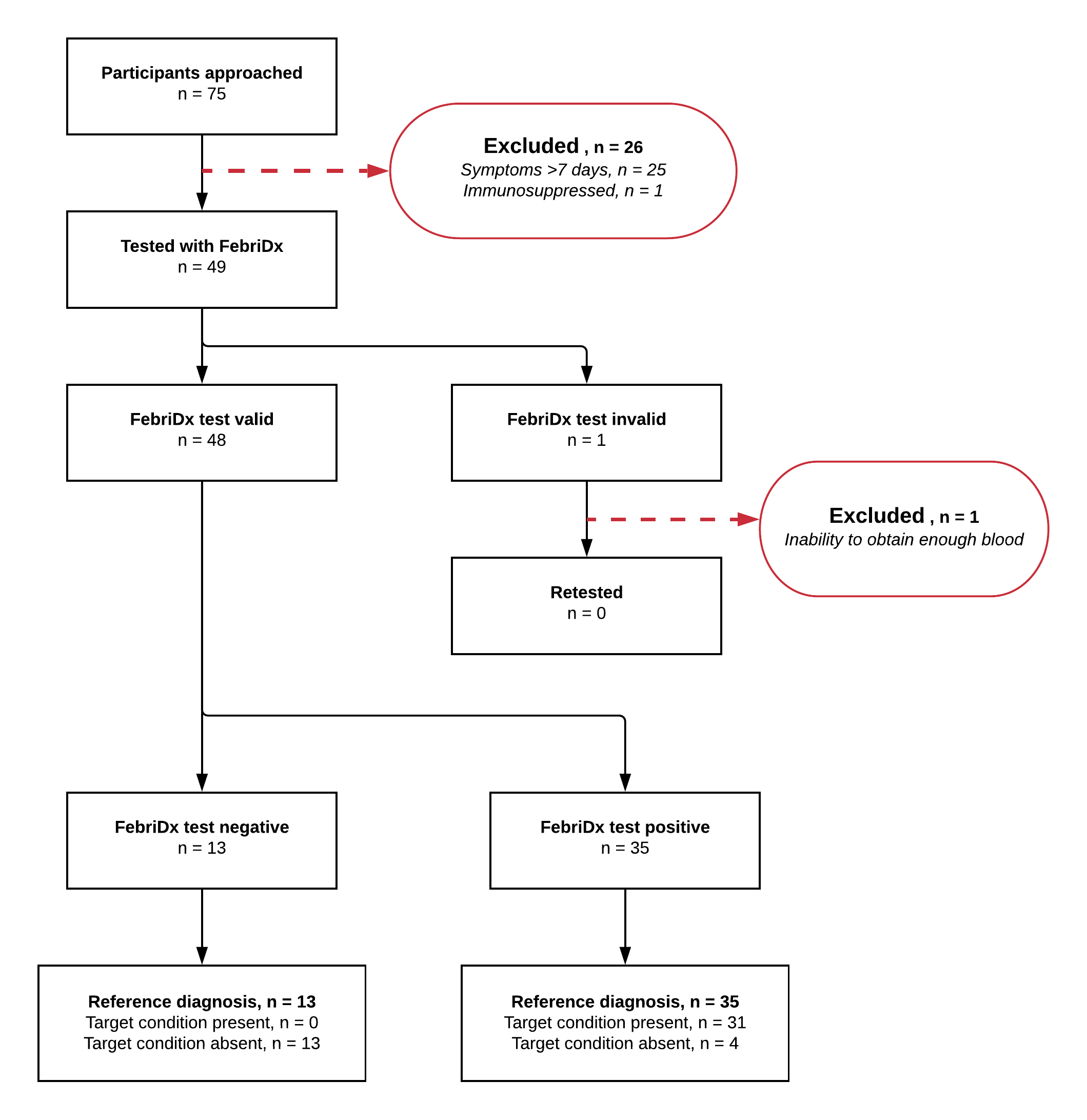

### Supplementary material 10

**Supplementary material 10**

The flow of patients in the London study (Houston et al.):

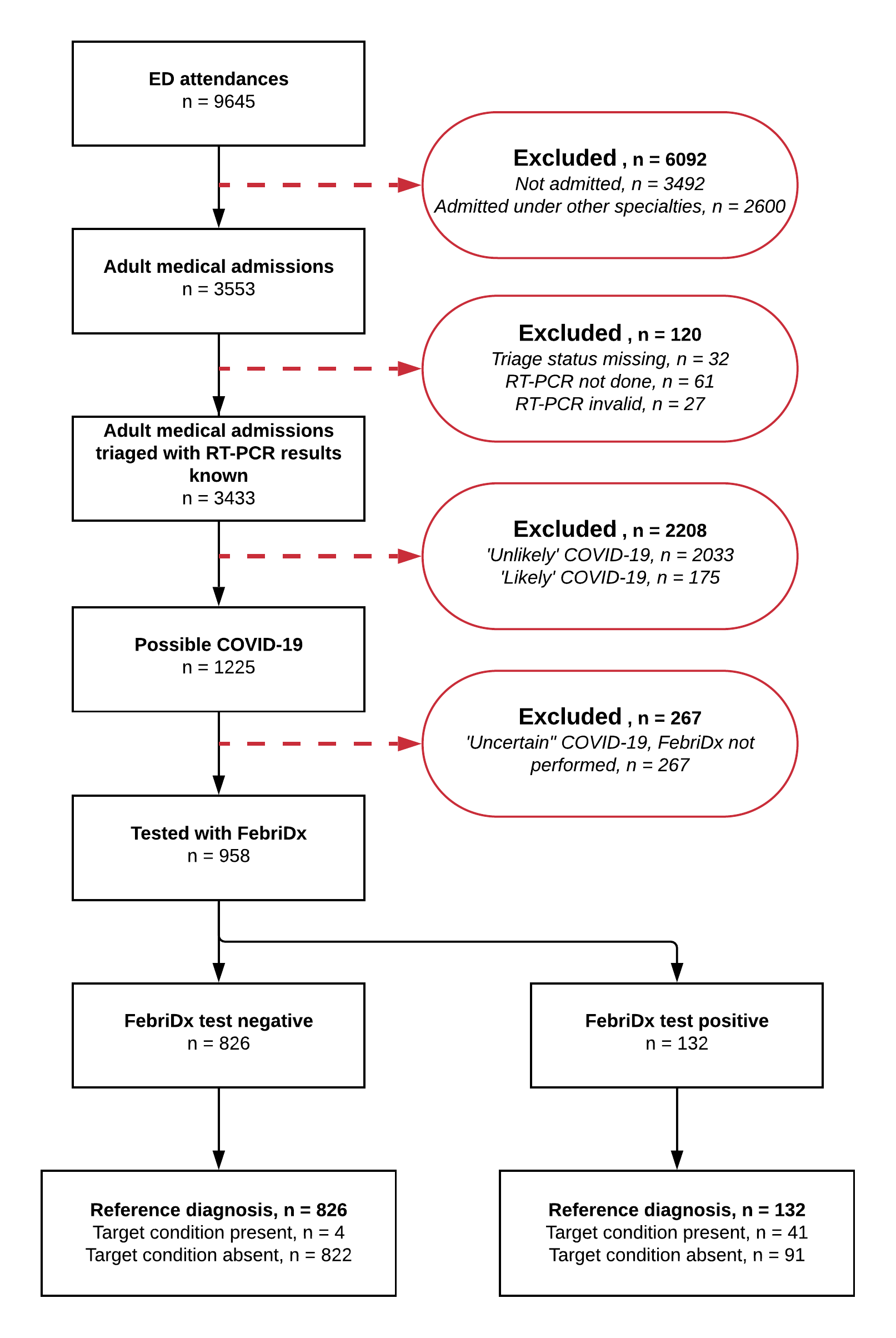
