## Supplementary material 7 for "FebriDx point-of-care test in patients with suspected COVID-19: a systematic review and individual patient data meta-analysis of diagnostic test accuracy studies"

Risk of bias (RoB) and applicability assessments for the included studies using the QUADAS-2 tool for the quality assessment of diagnostic accuracy studies. Signalling question 2, *“If a threshold was used, was it pre-specified?”*, from the ‘Index test’ domain was removed as it was not applicable to the review question.

These are summarised below for each of the four domains:

**Patient selection domain:**

|  | D1 Patient selection | | | | | | |
| --- | --- | --- | --- | --- | --- | --- | --- |
|  | A Risk of Bias | | | | | B Concerns regarding applicability | |
|  | Describe method of patient selection | Was a consecutive or random sample of patients enrolled? | Was a case-control design avoided? | Did the study avoid inappropriate exclusions? | Could the selection of patients have introduced bias? | Describe included patients | Is there concern that the included patients do not match the review question |
| **Southampton study** | Patients were prospectively enrolled from the first 266 patients consecutively approached for FebriDx testing alongside the nasopharyngeal swabs for RT-PCR testing.  Inappropriate exclusion may include 4/266 patients excluded for no having CRP line despite having a laboratory CRP of >20mg/L. However, some patients were re-tested, but it was unclear whether these 4 patients were re-tested or not. | Yes | Yes | Yes | **Low risk** | Patients presenting with suspected COVID-19 +/- ARI were prospectively enrolled. Presentation, intended use and settings seem appropriate for this test. | **Low concern** |
| **Kettering study** | Patients were prospectively enrolled and offered FebriDx at the same time as the nasopharyngeal swab for RT-PCR testing when meeting PHE definition of COVID-19 and requiring admission for at least one night.  27/75 patients were excluded (25 due to symptoms longer than 7 days). The FebriDx instructions for use state that patients should only be tested that have symptoms in the last 7 days, therefore it is not inappropriate to exclude patients with symptoms longer than 7 days but this is unlikely to reflect clinical setting use and is a potential source of bias in the current review. | Yes | Yes | Yes | **High risk** | Patients requiring admission for at least one night with suspected COVID-19. Presentation, intended use and settings seem appropriate for this test. | **Low concern** |
| **London**  **study** | Patients were selected from data prospectively entered into a COVID-19 triage database and retrospective extraction of clinical and bed allocation data from electronic patient records and hospital IT systems. Patients requiring medical admission were triaged into three categories for their likelihood of COVID-19 (unlikely, possible and likely). Patients in the possible group underwent testing with FebriDx. All patients received a nasal/pharyngeal swab for RT-PCR testing.  267/1225 patients were excluded, 145 due to unclear reason why the patient was not tested with FebriDx, which is a potential source of bias in the current review. | Yes | Yes | No | **High risk** | Patients requiring admission to a medical ward from the Emergency Department. Presentation, intended use and settings seem appropriate for this test. | **Low concern** |

**Index test domain:**

|  | D2 Index Test | | | |
| --- | --- | --- | --- | --- |
|  | A Risk of bias | | | B Concerns regarding applicability |
|  | Describe the index test and how it was conducted and interpreted | Were the index test results interpreted without results of the reference standard? | Could the conduct or interpretation of the index test have introduced bias? | Is there concern that the index test, its conduct, or interpretation differ from the review question? |
| **Southampton study** | FebriDx was conducted at the same time at nasopharyngeal swabbing for RT-PCR testing. MxA was interpreted as viral infection regardless of CRP expression. The result was interpreted by two investigators with a third adjudicating any disagreements. Testing was conducted according to manufacturer guidelines. | Yes | **Low risk** | **Low concern** |
| **Kettering**  **study** | FebriDx was conducted at the same time at nasopharyngeal swabbing for RT-PCR testing. MxA was interpreted as viral infection regardless of CRP expression. CRP alone was regarded as bacterial infection. Any borderline results and all negative results were verified by two investigators. Testing was conducted according to manufacturer guidelines. | Yes | **Low risk** | **Low concern** |
| **London**  **study** | FebriDx was conducted at a similar time as nasopharyngeal swabbing for RT-PCR testing. MxA was interpreted as viral infection regardless of CRP expression (results from the CRP window were not used given all patients had laboratory CRP measurements). It is unknown how many investigators interpreted the result. | Yes | **Low risk** | **Low concern** |

**Reference standard domain:**

|  | D3 Reference Standard | | | | |
| --- | --- | --- | --- | --- | --- |
|  | A Risk of bias | | | | B Concerns regarding applicability |
|  | Describe the reference standard and how it was conducted and interpreted | Is the reference standard likely to correctly classify the target condition? | Were the reference standard results interpreted without the results of the index test? | Could the reference standard, its conduct, or its interpretation have introduced bias? | Is there concern that the target condition as defined by the reference standard does not match the review question? |
| **Southampton study** | RT-PCR by either QIAstat-Dx or PHE laboratory RdRp and envelope protein (E) gene PCR assays conducted and interpreted as per standard guidelines. Binary results were provided, minimising user interpretation. Any RT-PCR positive results were considered positive for COVID-19. The reference standard alone is typically not sufficient, but combined with the inclusion criteria, suspicion was high in RT-PCR positive patients | Yes | Yes | **Low risk** | **Low concern** |
| **Kettering study** | RT-PCR by PHE laboratory RdRp and envelope protein (E) assays. Interpretation is not described in detail but its conduct and results appear consistent with this test. The reference standard alone is typically not sufficient, but combined with the inclusion criteria, suspicion of COVID-19 was high in RT-PCR positive patients. | Yes | Yes | **Low risk** | **Low concern** |
| **London**  **study** | RT-PCR by Panther Fusion SARSCoV-2, Abbott RealTime SARS-CoV-2, or an extraction free SARS-CoV-2 RT-PCR assay developed by Health Services Laboratories. Rapid RT-PCR assays were also used, Xpert Xpress SARS-CoV-2 or SAMBA II SARS-CoV-2, but these were prioritised for a different group of patients to those included in the current review. The reference standard alone is typically not sufficient, but combined with the inclusion criteria, suspicion was moderate in RT-PCR positive patients included in the current review. | Yes | Yes | **Low risk** | **Low concern** |

**Flow and timing domain:**

|  | D4 Flow and Timing | | | | | | |
| --- | --- | --- | --- | --- | --- | --- | --- |
|  | A risk of bias | | | | | | |
|  | Describe any patients who did not receive the index test(s) and/or reference standard or who were excluded from the 2x2 table | Describe the time interval and any interventions between index test(s) and reference standard: | Was there an appropriate interval between index test(s) and reference standard? | Did all patients receive a reference standard? | Did patients receive the same reference standard? | Were all patients included in the analysis? | **Could the patient flow have introduced bias?** |
| **Southampton study** | 18/266 patients were excluded. 15 due to declining consent or consultee declining assent, and 3 due to the inability to re-test them with FebriDx following an initial failure due to their clinical frailty. | Performed at the same time | Yes | Yes | Yes | No | **Low risk** |
| **Kettering study** | 27/75 patients were excluded. 25 due to having symptoms longer than 7 days, 1 due to being immunosuppressed, and 1 due to the inability to draw blood (re-testing was not deemed appropriate due to their clinical frailty). 25/75 patients excluded due to having symptoms longer than 7 days is a high proportion, although the FebriDx instructions for use state that patients should only be tested that have symptoms in the last 7 days, but this is unlikely to reflect clinical setting use and is a potential source of bias in the current review. | Performed at the same time | Yes | Yes | Yes | No | **High risk** |
| **London**  **study** | 267/1225 patients were excluded. 145 due to an unclear reason why they weren’t tested, 60 due to being immunosuppressed, 27 due to needing a high level of care, 20 due to having symptoms longer than 10 days, 13 due to previously having COVID-19, 1 due to being unable to bleed, and 1 refused testing with FebriDx. 145/1225 patients excluded due to an unclear reason why they weren’t tested is a high proportion and is a potential source of bias in the current review. | Performed at a similar time | Yes | Yes | Yes | No | **High risk** |

**Graphical summary of risk of bias results:**

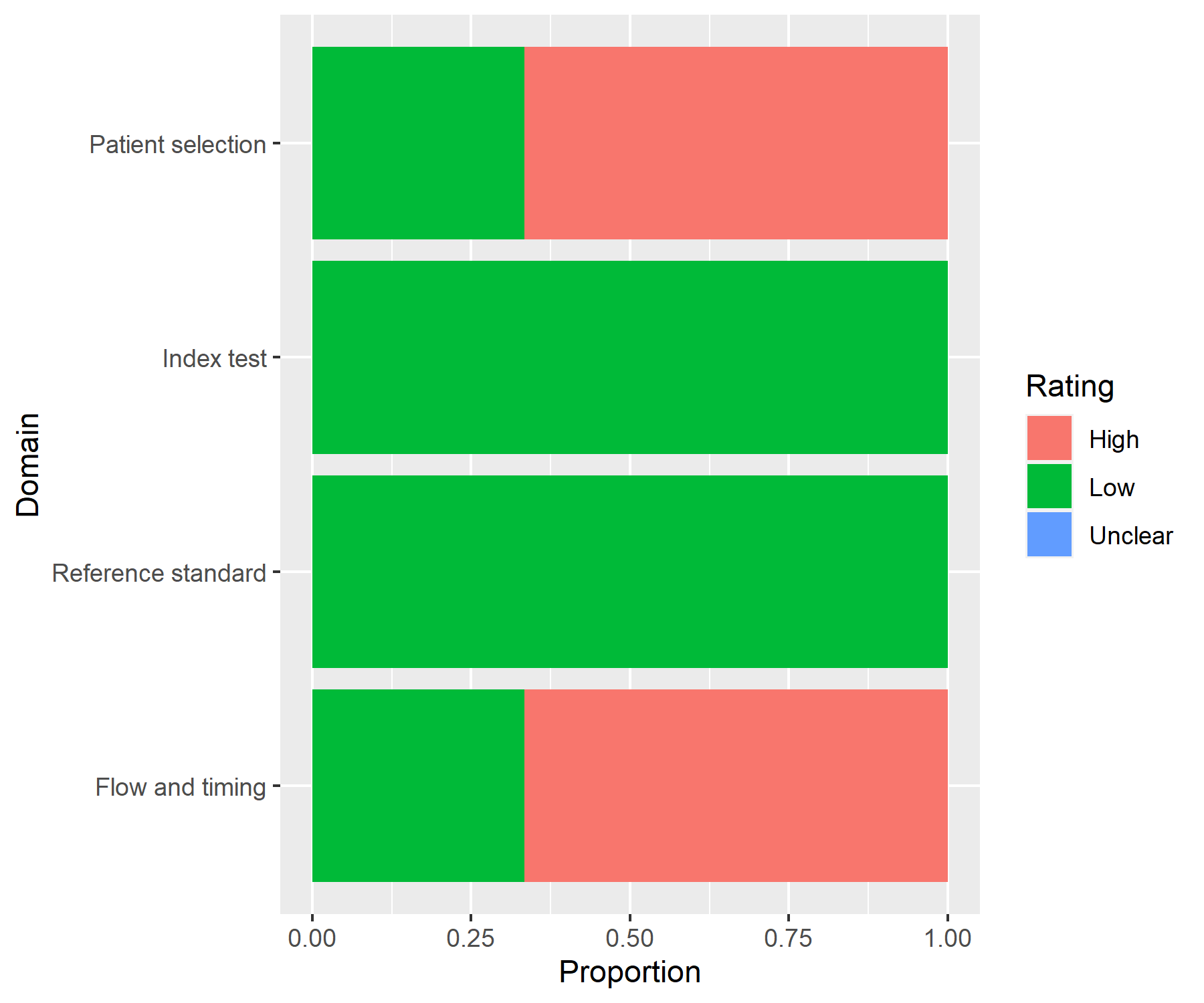

**Graphical summary of applicability results:**

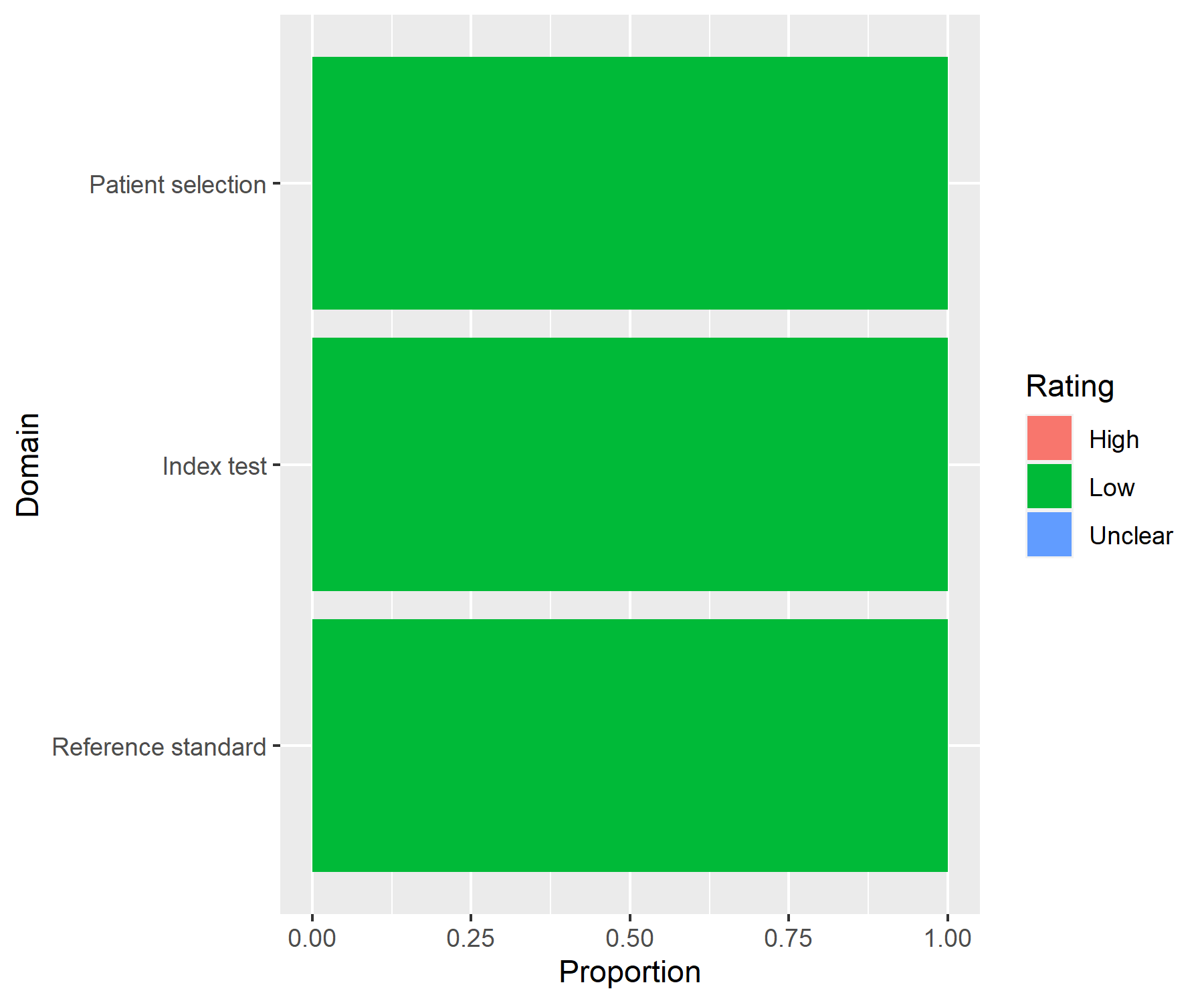
