## Supplementary material 8 for "FebriDx point-of-care test in patients with suspected COVID-19: a systematic review and individual patient data meta-analysis of diagnostic test accuracy studies"

The flow of patients in the Southampton study (Clark et al. + additional unpublished data):

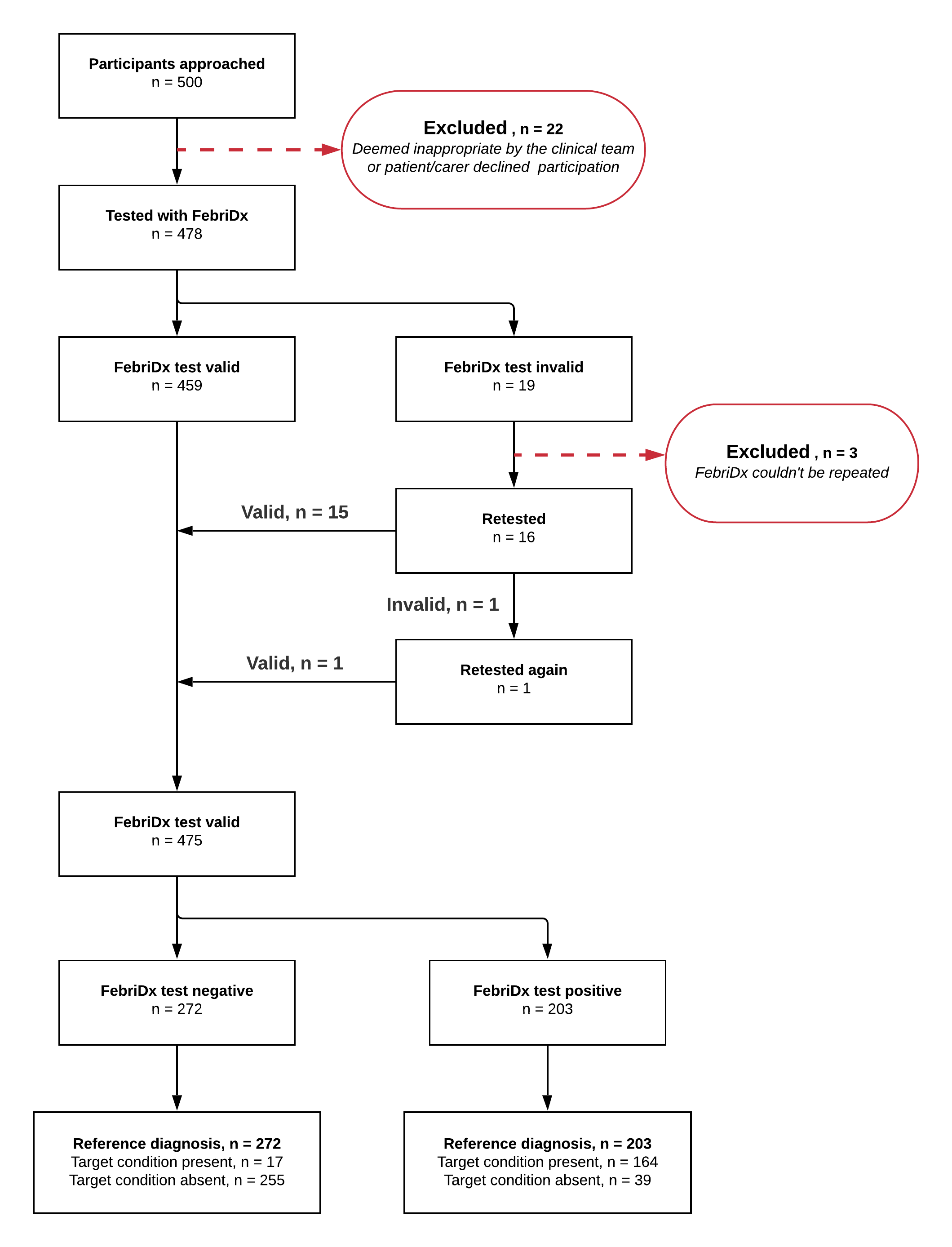
