## Supplementary material 11 for "FebriDx point-of-care test in patients with suspected COVID-19: a systematic review and individual patient data meta-analysis of diagnostic test accuracy studies"

The 2x2 contingency tables from the Southampton study, Kettering study, and London study for the symptom duration and age sub-groups.

**Southampton**

| Southampton – Symptom duration (0 to 7 days) | | | |
| --- | --- | --- | --- |
| *n =* | RT-PCR + | RT-PCR - | Total |
| FebriDx + | 105 | 25 | 130 |
| FebriDx - | 10 | 185 | 195 |
| Total | 115 | 210 | 325 |
| Southampton – Symptom duration (>7 days) | | | |
| *n =* | RT-PCR + | RT-PCR - | Total |
| FebriDx + | 59 | 14 | 73 |
| FebriDx - | 7 | 70 | 77 |
| Total | 66 | 84 | 150 |
| Southampton – Age (16-73 years) | | | |
| *n =* | RT-PCR + | RT-PCR - | Total |
| FebriDx + | 106 | 21 | 127 |
| FebriDx - | 11 | 151 | 162 |
| Total | 117 | 172 | 289 |
| Southampton – Age (>73 years) | | | |
| *n =* | RT-PCR + | RT-PCR - | Total |
| FebriDx + | 58 | 18 | 76 |
| FebriDx - | 6 | 104 | 110 |
| Total | 64 | 122 | 186 |

**Kettering**

| Kettering – Symptom duration (0 to 7 days) | | | |
| --- | --- | --- | --- |
| *n =* | RT-PCR + | RT-PCR - | Total |
| FebriDx + | 31 | 4 | 35 |
| FebriDx - | 0 | 12 | 12 |
| Total | 31 | 16 | 47 |
| Kettering – Symptom duration (>7 days) | | | |
| *n =* | RT-PCR + | RT-PCR - | Total |
| FebriDx + | 0 | 0 | 0 |
| FebriDx - | 0 | 0 | 0 |
| Total | 0 | 0 | 0 |
| Kettering – Age (16-73 years) | | | |
| *n =* | RT-PCR + | RT-PCR - | Total |
| FebriDx + | 22 | 3 | 25 |
| FebriDx - | 0 | 5 | 5 |
| Total | 22 | 8 | 30 |
| Kettering – Age (>73 years) | | | |
| *n =* | RT-PCR + | RT-PCR - | Total |
| FebriDx + | 9 | 1 | 10 |
| FebriDx - | 0 | 8 | 8 |
| Total | 9 | 9 | 18 |

**London**

| London – Symptom duration (0 to 7 days) | | | |
| --- | --- | --- | --- |
| *n =* | RT-PCR + | RT-PCR - | Total |
| FebriDx + | 29 | 74 | 103 |
| FebriDx - | 4 | 660 | 664 |
| Total | 33 | 734 | 767 |
| London – Symptom duration (>7 days) | | | |
| *n =* | RT-PCR + | RT-PCR - | Total |
| FebriDx + | 8 | 3 | 11 |
| FebriDx - | 0 | 52 | 52 |
| Total | 8 | 55 | 63 |
| London – Age (16-73 years) | | | |
| *n =* | RT-PCR + | RT-PCR - | Total |
| FebriDx + | 29 | 48 | 77 |
| FebriDx - | 4 | 359 | 363 |
| Total | 33 | 407 | 440 |
| London – Age (>73 years) | | | |
| *n =* | RT-PCR + | RT-PCR - | Total |
| FebriDx + | 12 | 43 | 55 |
| FebriDx - | 0 | 463 | 463 |
| Total | 12 | 506 | 518 |
